# Acceptance of an Mpox Vaccine in the Democratic Republic of the Congo

**DOI:** 10.1101/2024.08.15.24311971

**Authors:** Skylar Petrichko, Jason Kindrachuk, Dalau Nkamba, Megan Halbrook, Sydney Merritt, Handdy Kalengi, Leonard Kamba, Michael Beya, Nicole A. Hoff, Christophe Luhata, Didine K. Kaba, Anne W. Rimoin

**Author notes:** Corresponding Author:* Anne W. Rimoin, PhD, MPH, Professor, Department of Epidemiology, Gordon-Levin Endowed Chair in Infectious Diseases and Public Health, Jonathan and Karin Fielding School of Public Health, University of California, Los Angeles 650 Charles E. Young Dr, CHS 41-275, Los Angeles, CA, 90095. Denotes co-first authors. These authors contributed equally to this article. Denotes co-last authors (Didine K. Kaba, Anne W. Rimoin).

## Abstract

We report general acceptance of an mpox vaccine (61.0%) in the Democratic Republic of the Congo (n = 5226) with higher acceptance among healthcare workers and respondents in historic mpox-endemic regions. These data highlight the need for increased community engagement and sensitization before widespread deployment of mpox vaccines.

## INTRODUCTION

Mpox is a zoonotic infectious disease endemic to the Democratic Republic of the Congo (DRC) (1-3). In 2022, rapid spread of MPXV Clade IIb worldwide resulted in over 91,000 confirmed infections and the declaration of a public health emergency of international concern (PHEIC) (4). In the DRC incident cases have quadrupled between 2021 and 2023: over 28,000 suspected cases were reported from January 2023 to July 2024, and new introductions recorded in neighboring Burundi, Rwanda, Kenya, and Uganda (5). In August 2024 a travel-associated Clade Ib case was reported for the first time in Sweden (6). In response to the recent expanding burden of mpox, Africa CDC and the WHO declared a Public Health Emergency of Continental Security and second PHEIC, respectively.

Despite the deployment of a modified vaccinia Ankara-Bavarian Nordic vaccine to many high and middle-income countries, there currently is no mpox vaccine access for the general population in Africa. One vaccination model suggests that vaccination of 80% of children under 15 years old in DRC would result in robust reductions on morbidity, mortality, and mpox circulation (7).

## METHODS

The data for this cross-sectional analysis was drawn from a larger longitudinal phone survey which began in 2022 and included participants, targeting primarily HCWs, from all 26 provinces in the DRC. Participants were selected from historical phone survey records of over 10,000 persons. From these records, 5,226 adult participants were contacted between December 2023 to February 2024 by study staff to complete to this vaccine survey. Surveys were administered via phone by trained interviewers in Swahili, French, Lingala, Kikongo, and/or Tshiluba. Survey questions included behavioral and social drivers of vaccination and attitudes towards the introduction of new vaccines, including mpox. Mpox vaccine acceptance was measured by asking participants about their interest in the inclusion of a an mpox vaccine in the national vaccination schedule and participants were given several response choices—yes, for either adults, children or both; no; and I do not know this disease (Supplementary Material 1). All ‘yes’ responses were collapsed for analysis. Attitudes, vaccine acceptance by province, and sociodemographic characteristics were assessed via Likert scale questions and presented as a 5- or 3-level variable and chi-square test (α=0.05).

Ethics approval was provided by the Kinshasa School of Public Health (ESP/CE/188/2023) and the University of California, Los Angeles, as a public health response activity designation.

## RESULTS

Respondents were predominantly male (79.2%), with the highest percentage from Kwilu province (12.6%). There was roughly equivalent representation of those from urban or rural settings. Most respondents reported some college/university education (41.2%) and identified as healthcare workers (55.5%) **(Table 1)**.

**Table 1.**
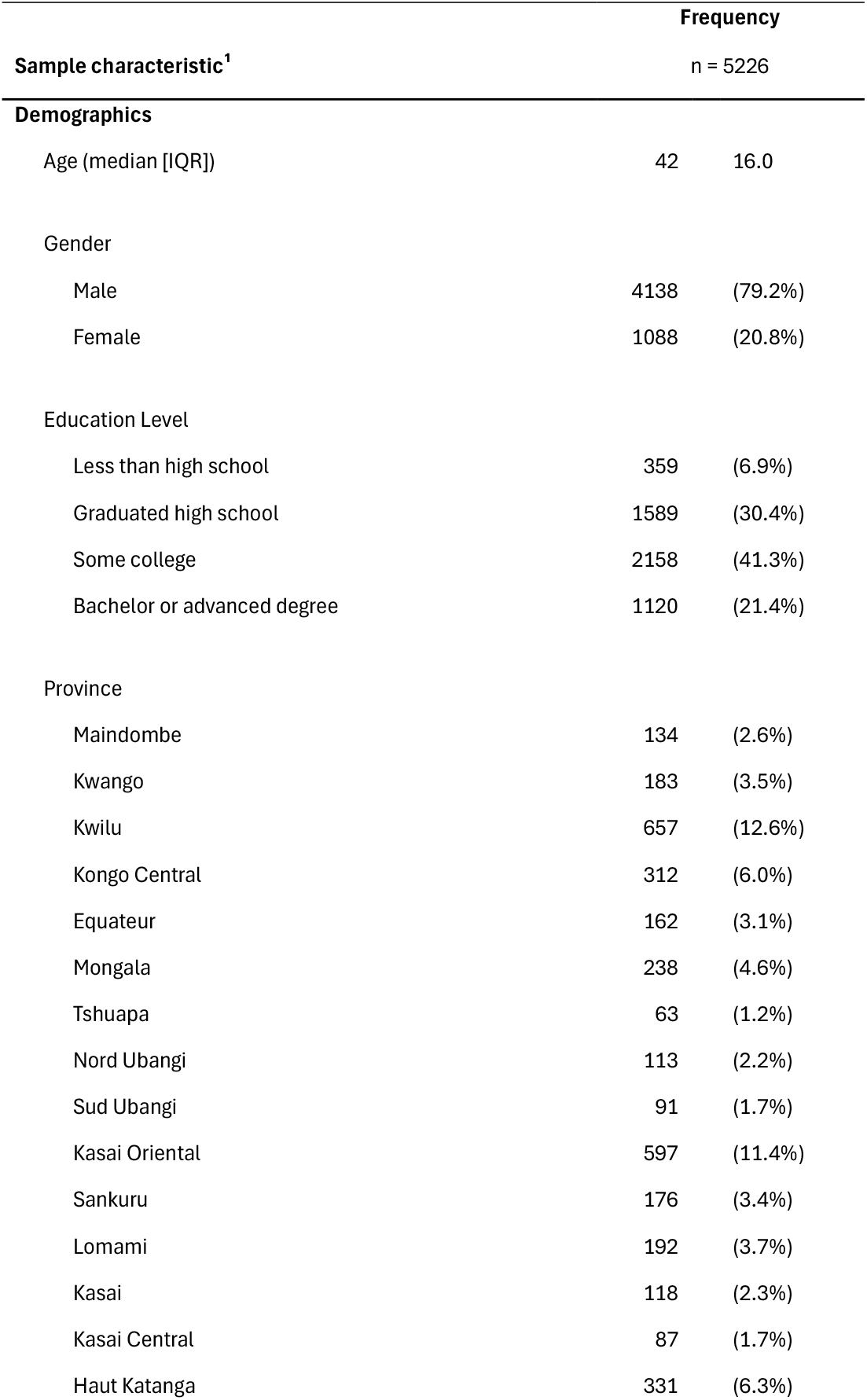
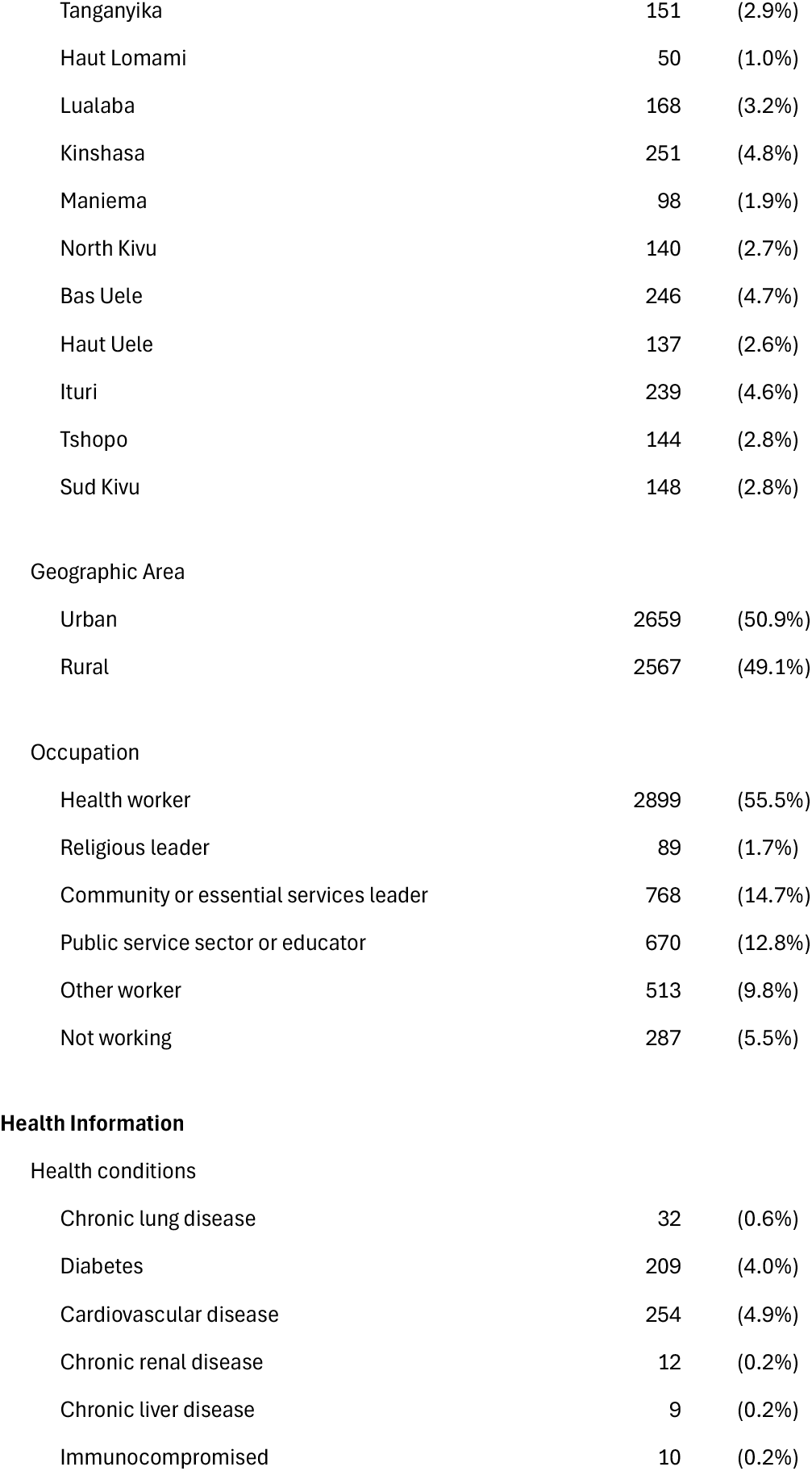
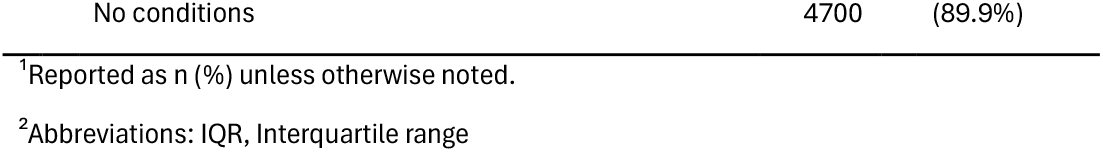
Demographic Characteristics of National Phone Survey Respondents (n = 5226)

Participants generally disagreed with the statement that new vaccines carry more risks than older vaccines. However, in four provinces—Sud Kivu, Nord Ubangi, Nord Kivu, and Lualaba—more than 50% of respondents agreed with or felt that greater risks exist. Respondents generally agreed with the statement that information received about vaccines from the vaccination program was reliable and trustworthy. In Sankuru province, 29.5% of respondents felt neutral about the reliability of vaccine program information; Haut Katanga and Lomami had the highest prevalence of vaccine information distrust, with 7.6% and 6.8% disagreeing with the statement. **(Figure 1)**.

**Figure 1.**
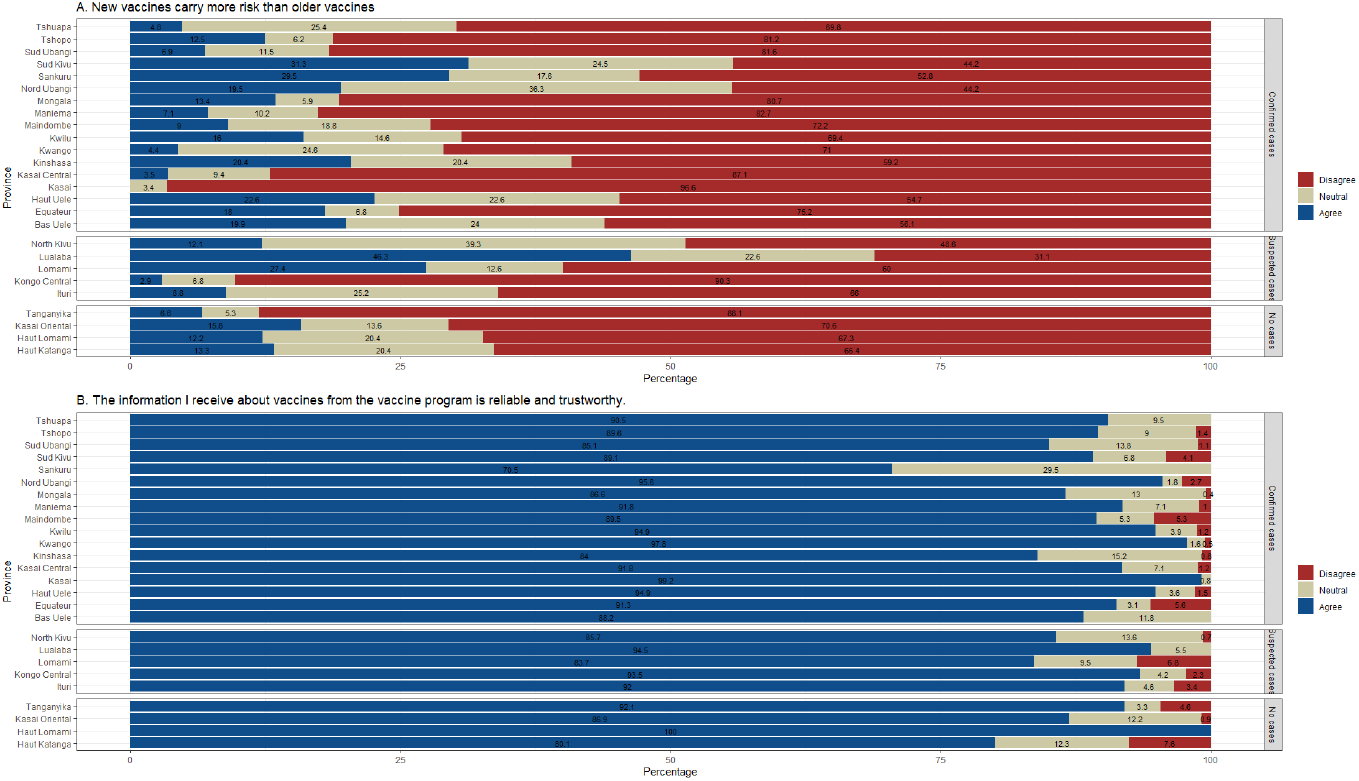
General Vaccine Attitudes and Perceptions by Province. (A) Reported responses to the statement “New vaccines carry more risks than older vaccines” (B) Reported responses to the statement “Information I receive about vaccines from the vaccine program is reliable and trustworthy”. 3-level responses were collapsed from a 5-point Likert scale.

Nationally, there was general acceptance of an mpox vaccination by all respondents (61.0%, 95% CI:59.6-62.4); 21.7% had no interest, and 17.3% reported no knowledge of mpox. At the provincial level, more than 80% of respondents from Kwango, Tshuapa, Nord Ubangi, Tanganyika, and Maniema were accepting of a vaccine, while respondents from Sankuru had the lowest acceptance (19.9%, 95% CI:14.3-26.6). Sankuru also had the highest percentage of participants that reported no mpox knowledge (57.4%, 95% CI:49.7-64.8). In general, rates of mpox vaccine acceptance and mpox disease knowledge were not influenced by the history of mpox cases within the province **(Figure 2)**.

**Figure 2.**
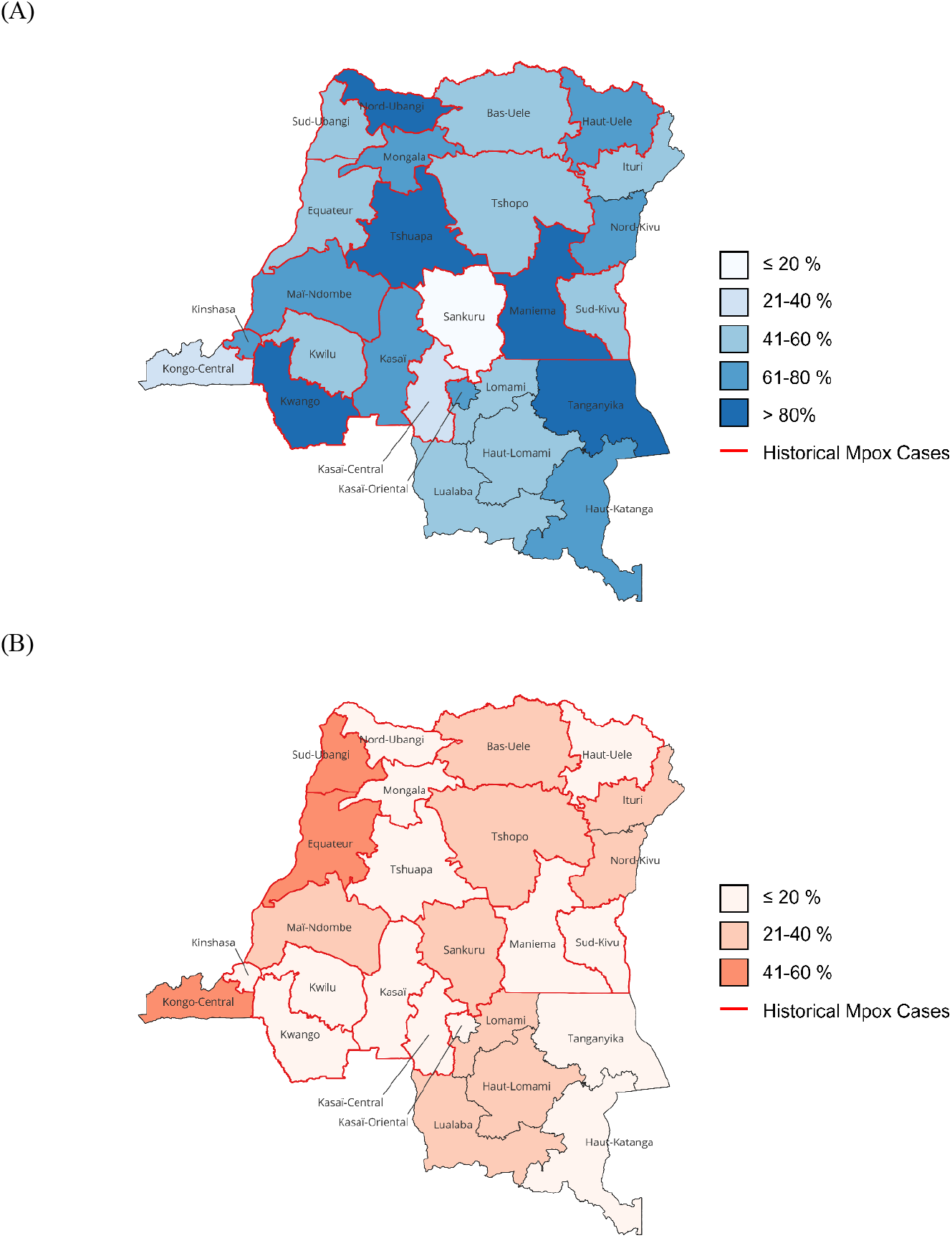
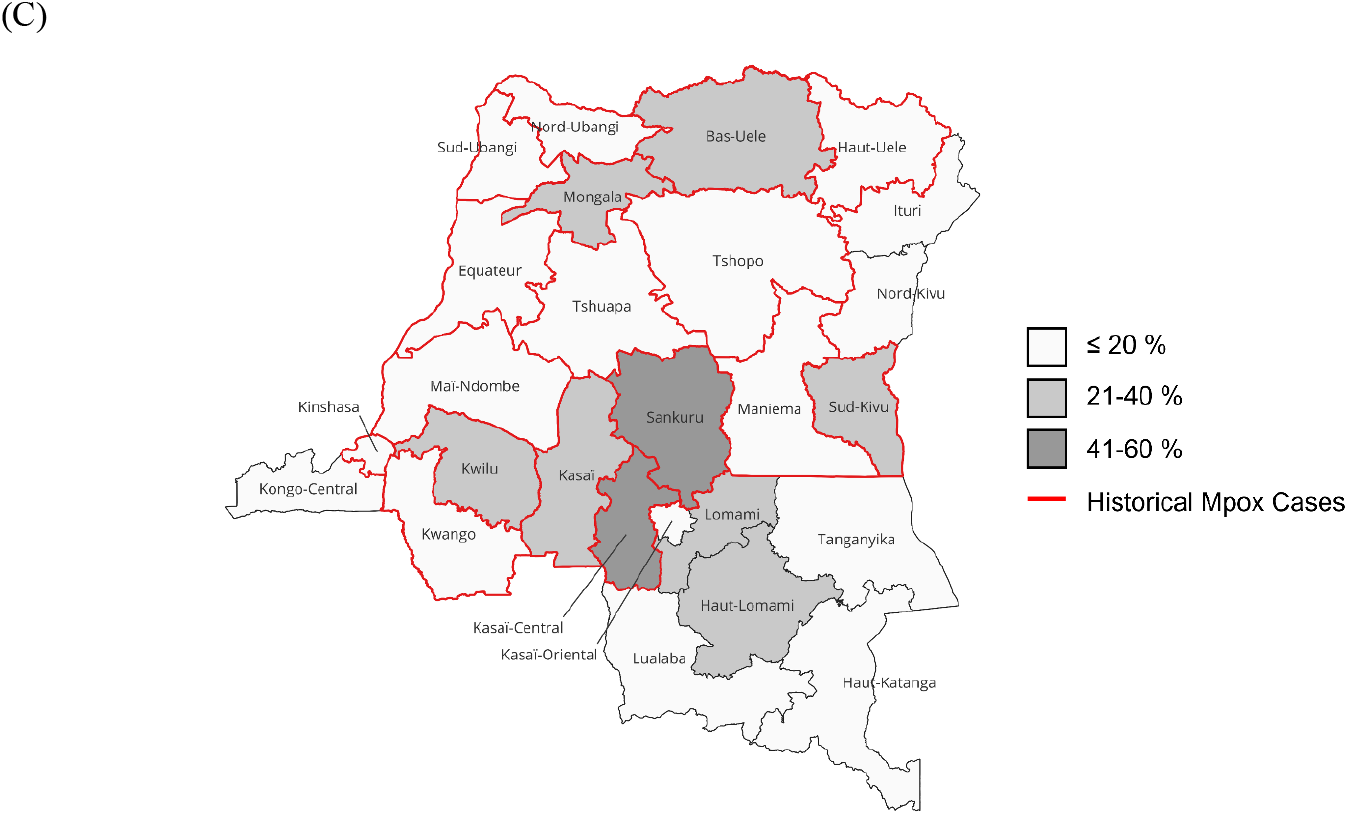
Map of Overall Mpox Acceptance by Province. (A) Reported acceptance as the collapsed responses of “Yes, for all populations” and “Yes, for children only” and “Yes, for adults only” (B) Reported non-acceptance by the response “No, not interested”. (C) Reported lack of knowledge of mpox disease. Percentages are reported by province.

Mpox vaccine acceptance varied significantly by province (p <0.0001). When stratified by educational attainment, those with a high school diploma had significantly lower acceptance of the mpox vaccine (49.8% 95%CI: 47.3-52.3) compared with others. Of all occupations reported, healthcare workers had the highest acceptance rates (69.4%, 95% CI: 67.6-71.1). Respondents from rural locations indicated greater acceptance of mpox vaccine deployment than their urban counterparts (64.4% vs 57.7%, p<0.0001). Among respondents who reported receiving a COVID-19 vaccine (n=4396), 63.2% indicated mpox vaccine acceptance. Additionally, persons who reported experiencing no chronic conditions reported higher rates of mpox vaccine acceptance (62.2% vs 51.0%) **(Figure 3)**.

**Figure 3.**
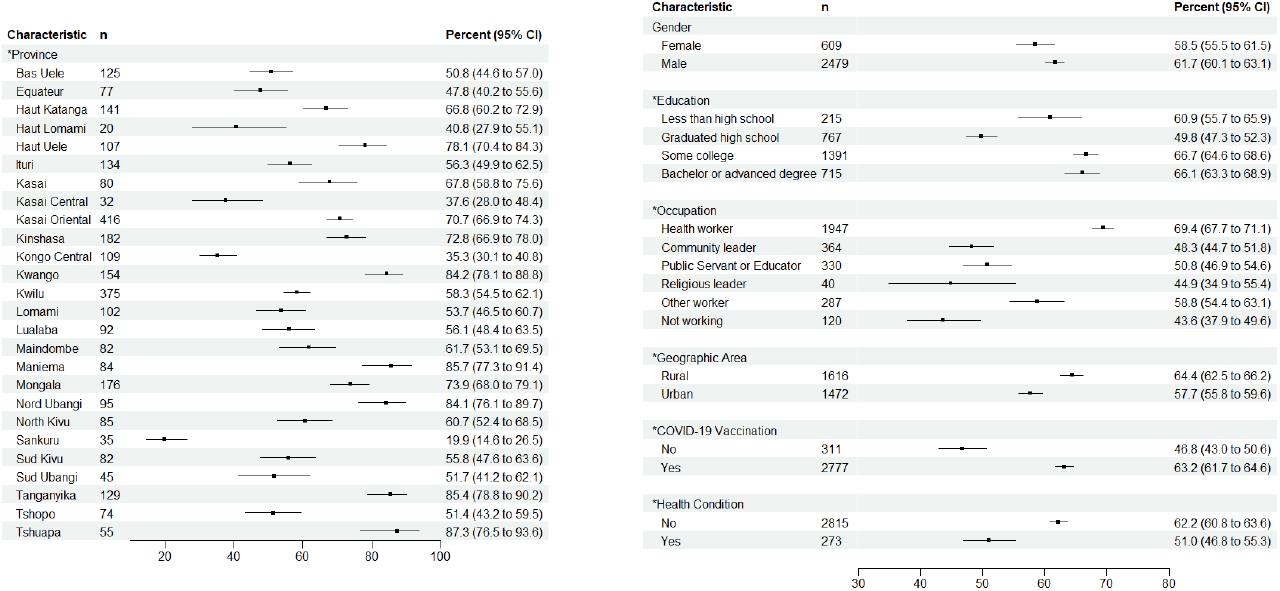
Percent of Mpox Vaccine Acceptance by Province and Demographic Characteristic. Acceptance of an Mpox Vaccine among respondents as stratified by sociodemographic characteristics and known mpox risk factors COVID-19 vaccination was dichotomized by whether the respondent received the COVID-19 vaccine. Health condition was dichotomized based on whether the respondent stated they had a chronic disease or were immunocompromised. ^*^ Chi-sq p-value <.05

## CONCLUSION

Given the ongoing escalation of the concurrent DRC Clade Ia and Clade Ib outbreaks and recent identification of cases in adjacent countries in Central Africa, there is increasing need for the rapid introduction of an mpox vaccine. However, to date, there is a paucity of information regarding mpox vaccine acceptance and mpox knowledge across DRC. We observed greater interest in mpox vaccine deployment among respondents from rural locations compared with urban populations. This is perhaps explained by the historic burden of mpox in DRC as mpox cases were primarily identified among children in rural regions. Currently, reported mpox cases are rising in urban centers, further challenging response and vaccine planning efforts.

Assessing mpox vaccine trust and acceptance across provinces that have and have not experienced mpox cases, and across demographic variables did not yield clear or neatly described trends. Instead, we observed several instances of outliers that may reflect the uniqueness and diversity of the DRC. For example, differing vaccine acceptance rates between those with and without chronic conditions may indicate underlying health anxieties among those with persistent health problems; further investigation is needed to better understand this relationship. Additionally, Sankuru province was consistently a vaccine hesitant outlier. Sankuru had the lowest proportion of respondents who trust information received from national vaccine programs and the lowest mpox vaccine acceptance, primarily because 57.4% of respondents stated no knowledge of mpox. In DRC, the majority of mpox endemic provinces consist of rainforest or forest-savannah mosaic geographies; the southeast provinces are primarily savanna and grassland and with no history of reported mpox cases. However, in June 2024 Lualaba reported their first mpox case. Interestingly, this province had the highest proportion of respondents believing that new vaccines have greater risks than older vaccines, though overall trust in national vaccine programs was high.

While these results report an overall trend of vaccine trust and acceptance nation-wide, they also indicate the need for a multifaceted approach to vaccine education and rollout. Strategies should be tailored to historically endemic regions and/or areas with lower vaccine acceptance, such as Sankuru or Kasai Central, and consider how personal experiences and cultural considerations intersect with mpox outbreaks. Of note, Haut Lomami province has been the setting of long-standing routine immunization revitalization efforts (8, 9), and 100% of respondents from this province reported that they feel information from the national vaccine program is reliable. This highly positive response is perhaps reflective of the many years of vaccine campaigns and education efforts in this region.

This survey was limited to those with access to phones; individuals with a lower socioeconomic status may have been inadvertently excluded from this study, including those in largely rural regions. This study included respondents from all provinces in DRC, with an emphasis on recruiting HCW. Past studies in the DRC have demonstrated 54% acceptance towards outbreak-related vaccines, with HCW continually expressing the highest acceptance among all respondents (10). As this study sought to enroll HCWs, and community and religious leaders, we observed an over-representation of men as they primarily hold these leadership roles in the DRC. As HCWs and religious leaders serve as health recommenders for their communities, there is a critical need to identify and address vaccine acceptance among these groups. These data provide insight regarding future deployments in adjacent regions as well as vaccine acceptance and deployment strategies within DRC, or regionally, for other emerging infectious diseases.

## Data Availability

All data produced in the present study are available upon reasonable request to the authors.

## ACKNOWLEDGEMENTS

All cartographic figures were generated using qGIS (Maidenhead, 3.36.3). We thank Brooke Aksnes and Emma Wray Aberle-Grasse for their continued support of this ongoing research endeavor.

